# COVID-19 Mortality Following Mass Gatherings

**DOI:** 10.1101/2020.10.27.20219717

**Authors:** Oren Miron, Kun-Hsing Yu, Rachel Wilf-Miron, Nadav Davidovitch

## Abstract

We examined Coronavirus Disease-2019 (COVID-19) mortality following 5 mass gatherings at outdoor rallies in the United States, during August 2020. We found that COVID-19 mortality started increasing 19-24 days after the mass gathering. In a 50-mile radius there was a 2.1-fold increase in COVID-19 mortality, and in a 51-100 miles radius there was a 1.4-fold increase. Our results suggest that precautions should be taken in mass gatherings and in at least a 50-mile radius, in order to limit COVID-19 mortality.

## Introduction

Mass gatherings have been linked to Coronavirus Disease-19 (COVID-19) deaths at the start of the pandemic, which led governments to temporarily limit most mass gatherings.^1^ This makes it difficult to determine the effect of mass gatherings on COVID-19 mortality in the later stage of the pandemic, which has more testing and treatments.^2^

If mass gatherings are followed by increased mortality, there is a need to estimate the radius of this effect in order to increase precautions in that region.

## Methods

Five mass gatherings occurred in August 2020 at outdoor rallies, in the states of Minnesota (August 17^th^), Wisconsin (August 17^th^), Arizona (August 18^th^), Pennsylvania (August 20^th^), and New-Hampshire (August 28^th^). We retrospectively extracted the daily COVID-19 mortality of each county in those states in the 30 days following each mass gathering.^3^

We aggregated the counties based on distance from their center coordinates to the coordinates of the mass gathering in their state, with groups of 0-50 miles, 51-100 miles, above 100 miles.^4^ We also extracted the mortality in all the states that did not have those mass gatherings in August 2020, and we defined their control mass gathering date as the median date of the August mass gatherings (August 18^th^).

We calculated the mortality rate per 100,000 capita with a 7-day moving-average, and examined its trend using the JoinPoint Regression Program (National Cancer Institute), and a 2-tailed statistical test with a significance level of 0.05. Lastly, we examined the rate and 95% confidence interval (CI) of the incidence when the gatherings are expected to have effects on deaths (20 days post-gathering)^5^, and compared it with the rate at 30 days post-gathering.

## Results

At 0-50 miles from the mass gathering, the COVID-19 death rate decreased from gathering day to day 24 (−1.6% daily-change, 95% confidence interval -2.4% to -0.9%), followed by an increase until day 30 (7.8% daily-change, 95% confidence interval 1.4% to 14.6%). At day 20 the rate was 0.07/100,000 capita (95% confidence interval, 0.04 to 0.1), and at day 30 it increased to 0.15/100,000 capita (95% confidence interval, 0.11 to 0.2, 2.1-fold increase; Figure 1).

**Figure 1:**
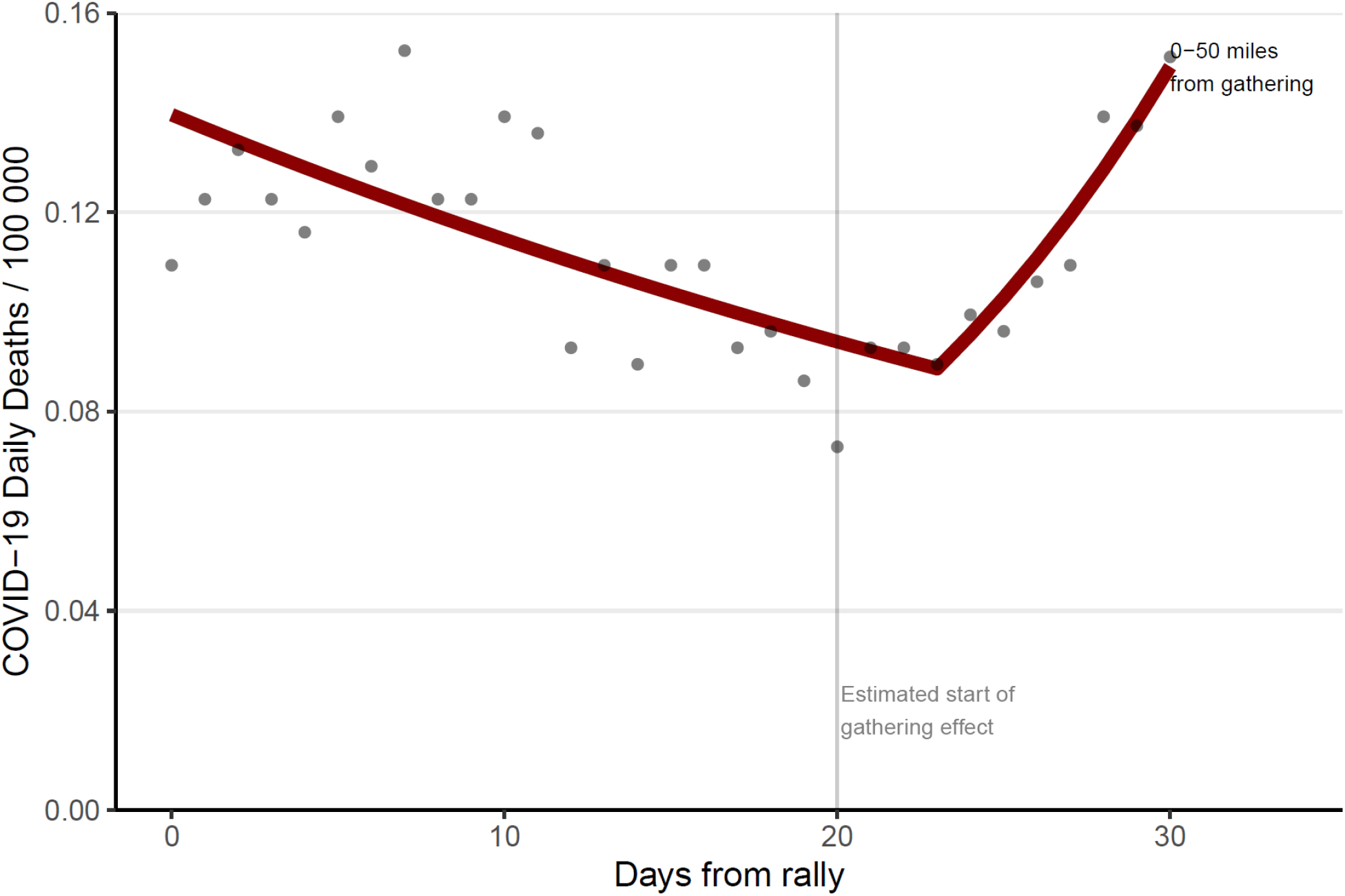
COVID-19 mortality by days in a 50 miles radius from mass gathering. Y indicates Coronavirus Disease-19 deaths per 100000 capita, and X indicates days from mass gathering. Dots indicate the observed data points from counties in the state of the mass gathering who were 50 miles or less from the gathering location. The line indicates the trend based on JoinPoint regression.

At 51-100 miles, the COVID-19 death rate decreased from gathering day to day 19 (−1.9% daily-change, 95% confidence interval -2.3% to -1.5%), followed by an increase until day 30 (3.9% daily-change, 95% confidence interval 2.9% to 4.8%). At day 20 the rate was 0.1/100,000 capita (95% confidence interval, 0.08 to 0.12), and at day 30 it increased to 0.15/100,000 capita (95% confidence interval, 0.12 to 0.17, 1.4-fold increase; Figure 2).

**Figure 2:**
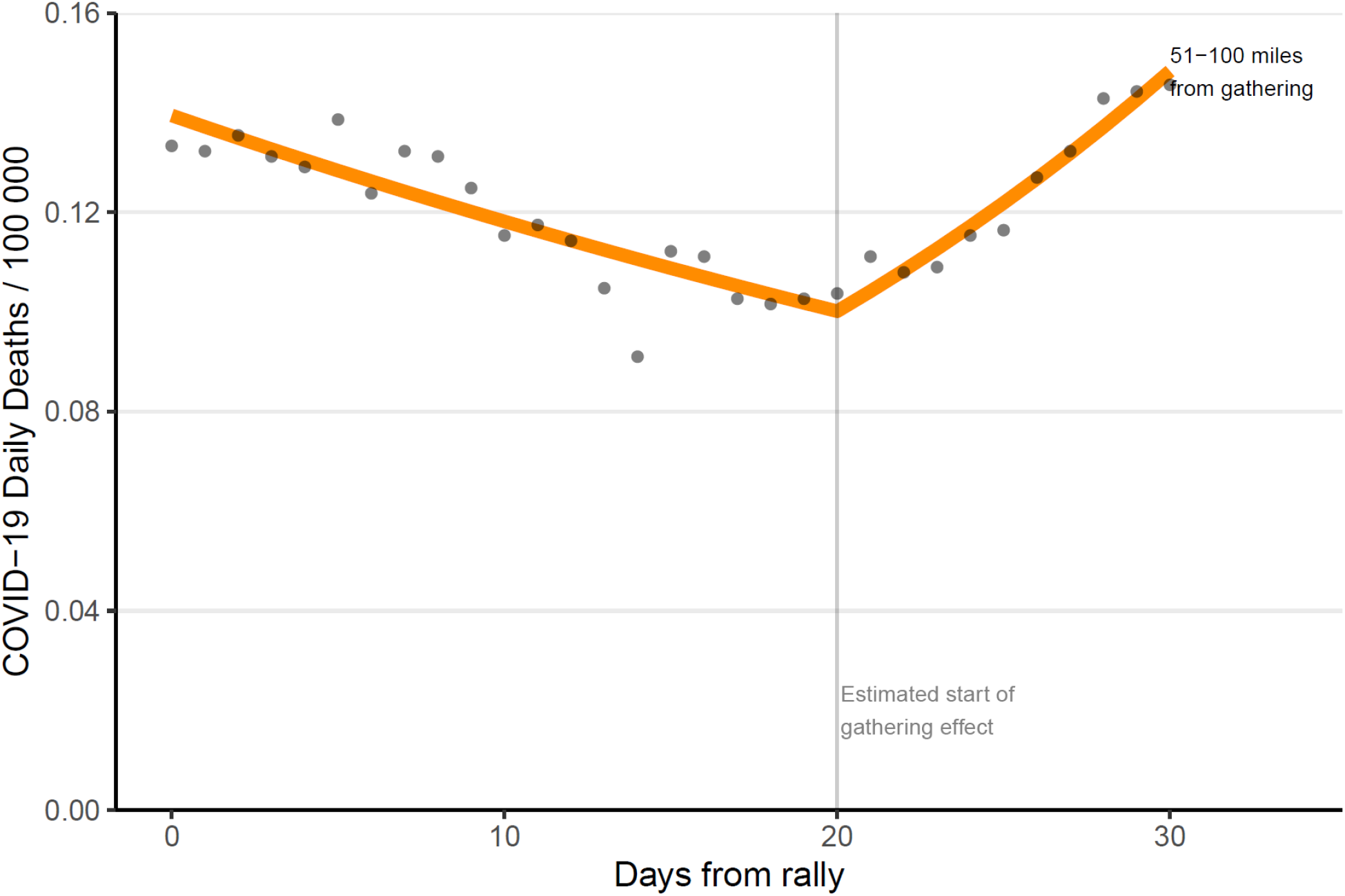
COVID-19 mortality by days in a 51-100 miles radius from mass gathering. Y indicates Coronavirus Disease-19 deaths per 100000 capita, and X indicates days from mass gathering. Dots indicate the observed data points from counties in the state of the mass gathering who were between 50-100 miles from the gathering location. The line indicates the trend based on JoinPoint regression.

At over 100 miles, the COVID-19 death rate decreased from gathering day to day 25 (−2.7% daily-change, 95% confidence interval -3.3% to -2.1%), followed by a daily change of 7% (95% confidence interval -0.4% to 15%). At day 20 the rate was 0.18/100,000 capita (95% confidence interval, 0.15 to 0.2), and at day 30 it increased to 0.22/100,000 capita (95% confidence interval, 0.2 to 0.25, 1.2-fold increase; Figure 3).

**Figure 3:**
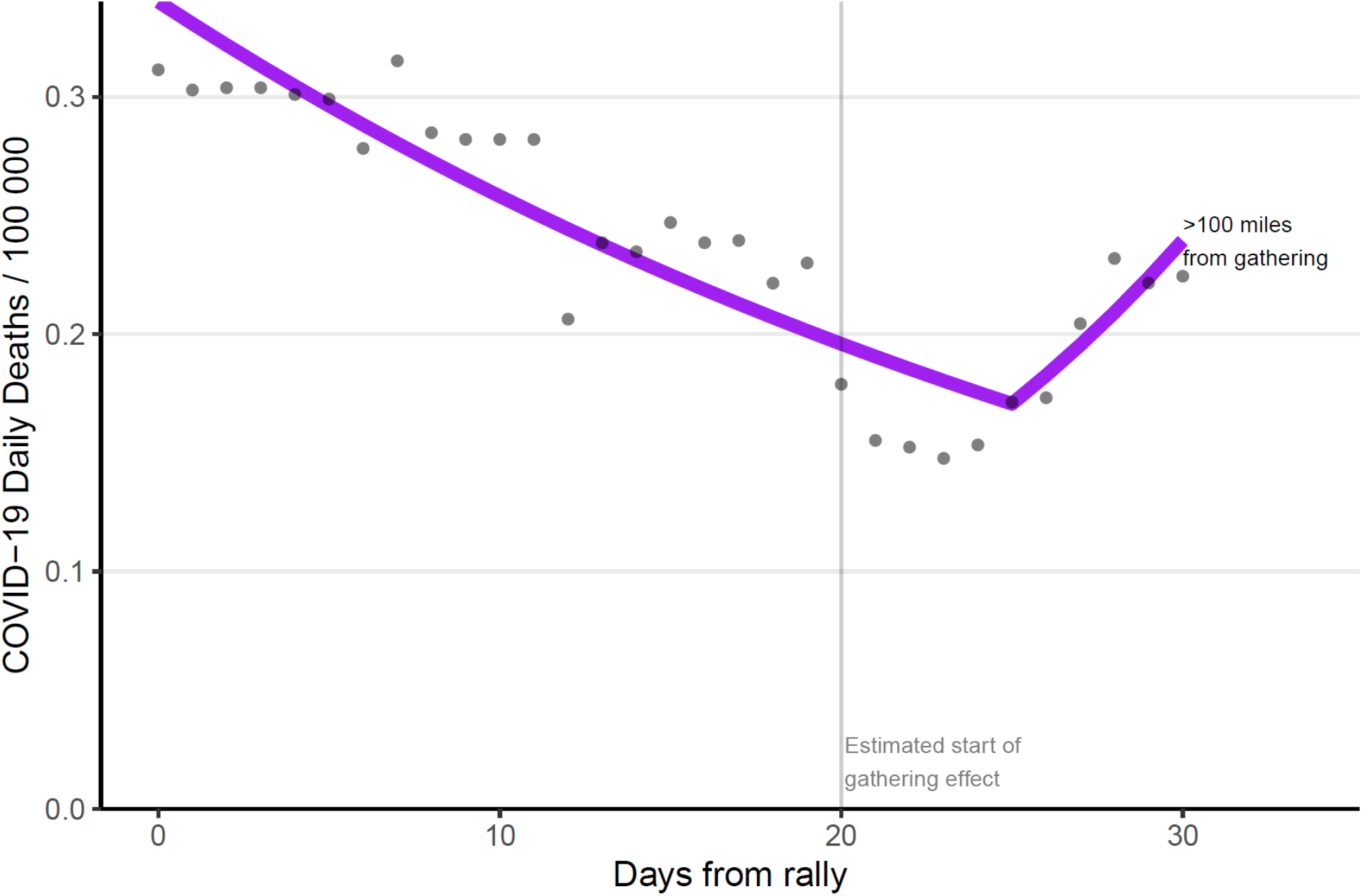
COVID-19 mortality by days in 100 miles or more radius from mass gathering. Y indicates Coronavirus Disease-19 deaths per 100000 capita, and X indicates days from mass gathering. Dots indicate the observed data points from counties in the state of the mass gathering who were over 100 miles from the gathering location. The line indicates the trend based on JoinPoint regression.

In states without the mass gatherings, the COVID-19 death rate decreased from control gathering day to day 20 (−1.1% daily-change, 95% confidence interval -1.4% to -0.8%), followed by an increase until day 30 (1.1% daily-change, 95% confidence interval 0.2% to 2.1%). At day 20 the rate was 0.27/100,000 capita (95% confidence interval, 0.26 to 0.27), and at day 30 it increased to 0.3/100,000 capita (95% confidence interval, 0.29 to 0.31, 1.1-fold increase; Figure 4).

**Figure 4:**
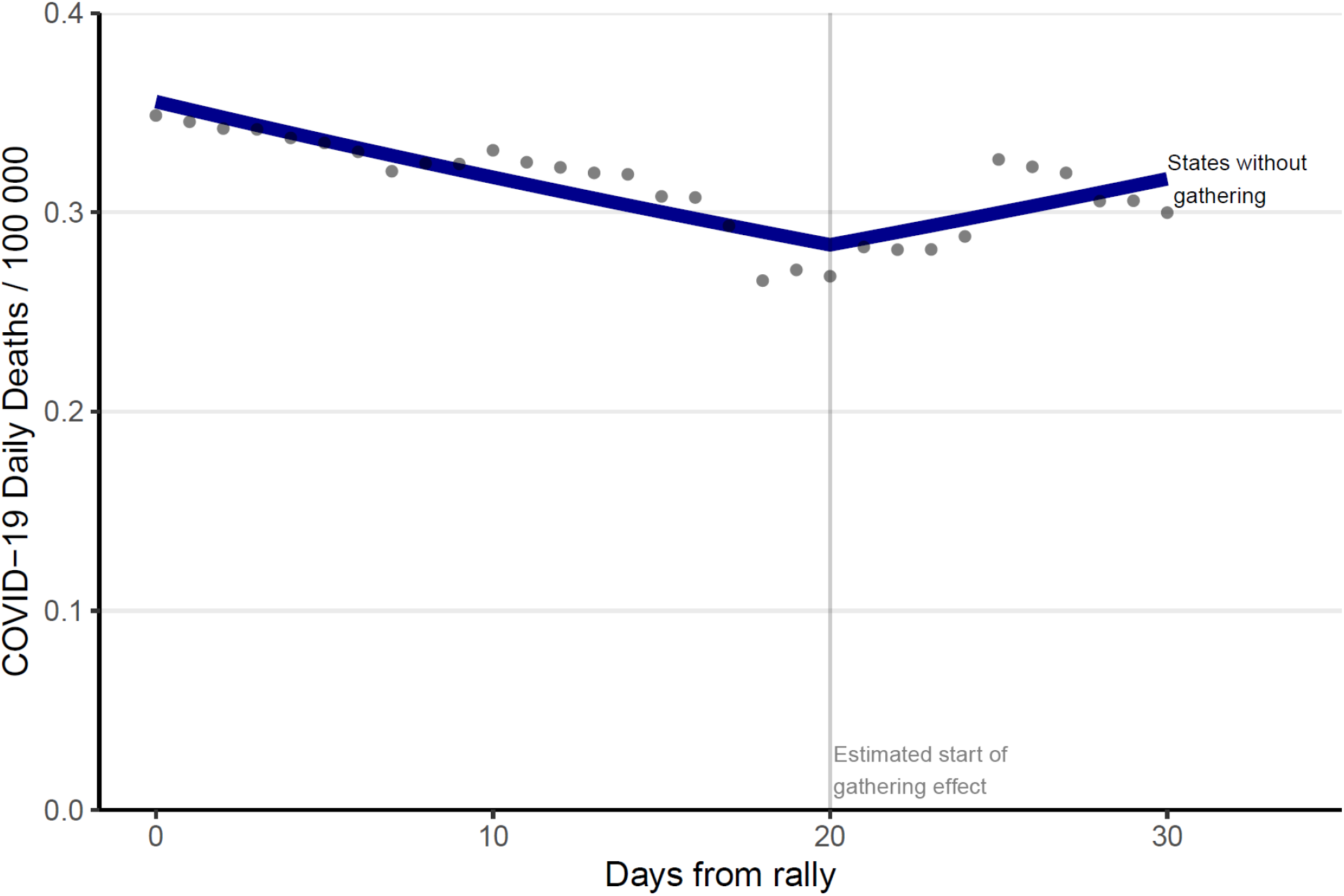
COVID-19 mortality by days from control date in states without mass gatherings. Y indicates Coronavirus Disease-19 deaths per 100000 capita, and X indicates days from mass gathering control date (median date of the 5 mass gatherings). Dots indicate the observed data points from counties in states who did not have any of the 5 mass gatherings that were analyzed. The line indicates the trend based on JoinPoint regression.

## Discussion

Our analysis showed an increase in COVID-19 mortality following mass gatherings, with the highest increase at 0-50 miles from the mass gathering (2.1-fold), followed by 51-100 miles (1.4-fold), 100+ miles (1.2-fold), and in states without the mass gatherings (1.1-fold).

The mortality increase in states without mass gatherings could stem from unrelated factors,^6,7^ or relate to out-of-state participants in the mass gatherings, such as the Georgia resident who died of COVID-19 after attending an Oklahoma mass gathering without a mask or distancing.^8^ States could promote masks and distancing in mass gatherings, and in at least a 50-mile radius from the mass gathering, which may limit COVID-19 mortality.

## Supporting information

STROBE checklist

## Data Availability

Publicly available

https://github.com/CSSEGISandData/COVID-19

## ARTICLE INFORMATION

### Author Contributions

Oren Miron had full access to all of the data in the study and takes responsibility for the integrity of the data and the accuracy of the data analysis.

Concept and design: All authors.

Acquisition, analysis, or interpretation of data: All authors. Drafting of the manuscript: Oren Miron.

Critical revision of the manuscript for important intellectual content: Yu, Wilf-Miron, and Davidovitch.

Statistical analysis: Oren Miron and Yu.

Study supervision: Davidovitch.

### Data availability statement

publicly available at https://github.com/CSSEGISandData/COVID-19

